# Predicting Huntington’s disease state with ensemble learning & sMRI: more than just the striatum

**DOI:** 10.1101/2023.07.24.23293076

**Authors:** Maitrei Kohli, Dorian Pustina, John H. Warner, Daniel C. Alexander, Rachael I. Scahill, Cristina Sampaio, Sarah J. Tabrizi, Peter A. Wijeratne

## Abstract

Developing effective treatments for Huntington’s disease (HD) requires reliable markers of disease progression. Striatal atrophy has been the hallmark of HD progression, but volumetric anomalies are also found in other brain regions. Little is known about the potential increase in predictive biomarking accuracy when volumetric scores from multiple brain regions are combined to predict the HD status of individual participants. We used cross-sectional structural MRI data from 184 HD gene-positive participants to a) test a novel ensemble machine learning model in classifying participants in one of four HD progression states (PreHD A; PreHD B; HD1; HD2), and (b) identify the brain regions that carry HD biomarking signal from 15 regions. We used 5-fold cross validation and backward feature elimination to find the optimal predictors and investigated the stability of the findings through repeated analyses. The ensemble predictive model systematically matched or outperformed the accuracy of nine standard machine learning models, reaching 55.3%±6.1 balanced accuracy in 4-group classification. The accuracy was higher for binary classifications (PreHD vs HD: 83.3%±6.3; PreHD A vs PreHD B: 76.7%±8.0; PreHD B vs HD1: 75.9%±8.5; HD1 vs HD2: 70.9%±9.4). Striatal structures (caudate and putamen) were systematically found to be top predictors. However, the accuracy increased substantially when we included other regions in the model (e.g., occipital cortex, lateral ventricles, cingulate, temporal lobe). Optimal models frequently included 2-7 brain regions from different areas. Overall, the accuracy of classifications remained stable across repetitions but the list of selected brain regions could vary, likely due to collinearities in volumetric scores. This is the first study to demonstrate the improvement of classification accuracy when predicting HD progression with a stacked ensemble model. Our findings indicate that HD progression is marked not only by striatal atrophy but also by volumetric changes outside the striatum, without which biomarking models cannot achieve optimal results. The robust methods applied here exposed instability in the selection of brain regions despite the sizeable sample size (n=184); such instabilities could lead to different conclusions in different studies when single analyses are applied on smaller sample sizes. From a translational perspective, our study informs on the selection of candidate endpoints or target regions for therapeutic intervention in future clinical trials.

## Introduction

Huntington’s disease (HD) is a monogenic, autosomal-dominant fatal neurodegenerative condition characterized by motor, cognitive, and behavioural symptoms.^1, 2^ The genetic marker for HD—an expansion of a CAG tract in the huntingtin gene to more than 39 repeats—is fully penetrant.^3, 4^ The first known neurodegenerative processes in HD are observed most notably in the striatum, beginning in the caudate, affecting mainly the medium spiny neurons, and progressing ventrally and laterally to the putamen^5^; degeneration in both the caudate and the putamen are hallmarks of HD neuropathology.^2, 5, 6^

Group studies in HD have shown substantial neurodegeneration at least a decade prior to the clinical motor diagnosis (CMD).^1, 7–9^ Volume loss as measured by structural MRI (sMRI) is one of the most studied biomarkers of HD.^39^ The gradual atrophy emerging many years before clinical manifestation indicates that therapeutic intervention may achieve maximal benefit when applied early to contain the degenerative process.^1, 7^ Despite much interest in the use of sMRI-derived measures as biomarkers, their impact on actual clinical practice has been limited. Scientific analyses are often conducted at the group-level whereas clinical practice requires biomarkers that can be applied at the individual-level; i.e., inclusion/exclusion criteria, patient stratification into groups, etc. For neuroimaging techniques such as sMRI to be useful in clinical settings, they must be able to make inferences at the level of the individual.^10^

Machine learning (ML) methods are widely employed when building data-driven models of disease state.^11^ The benefits of applying ML methods with sMRI data are twofold. First, ML methods allow characterisation at the individual level and are therefore more clinically translational. Second, given their multivariate nature, these approaches are sensitive to distributed and subtle effects in the brain that would otherwise be indiscernible using univariate methods that rely on differences in individual brain regions.^10^ Some studies that have used neuroimaging and machine learning for disease state classification in HD are summarized in supplementary material section S1.

Despite a growing interest in exploring ML methods, studies (supplementary material section S1) have mostly focused on binary classification problems, such as discriminating between healthy controls (HC) and HD, or premanifest (PreHD) and HC. However, since diagnosis can be confirmed by genetic testing, these models add little value. There is therefore a pressing need to better characterise the volumetric changes in the premanifest phase to identify the most suitable candidates for therapeutic intervention. What is more difficult but more useful would be to discern between PreHD vs HD or even more finely, far from CMD (PreHD A) vs close to CMD (PreHD B) with a view to predicting those likely to be approaching clinical onset.

Here, we address this issue starting with testing ML models on a wider spectrum of disease progression and classifying participants in finer disease states. Moreover, previous studies have often relied on small sample sizes, which may hamper the ability to build robust, unbiased predictive models that generalise to the HD population. Lastly, most methods preselect the features or identify regions-of-interest a priori, which works well for building an optimal predictive model but it is not suitable when applied for knowledge discovery; rather, it might mislead. For instance, if several sets of risk factors are equally predictive of an event, then it is misleading to return only one of them and disregard the rest.

Here we present a stacked ensemble-based ML model for the predictive classification of individual-level HD states that is robust to the common methodological challenges discussed above. Volumetric measures derived from sMRI enable the identification of HD-related brain alterations.^21^ ML models such as stacked ensemble^22^ allow data-driven individual-specific predictions of disease state.^5^ Quantifying feature importance aids interpretation of model outcomes by identifying which brain regions carry most discriminative information. We used baseline cross-sectional sMRI data from 184 HD gene-positive participants from the TRACK-HD^3^ dataset to (a) classify HD state using the 2-tier stacked ensemble ML model, and (b) identify which brain regions (i.e., features) carry most discriminative information for each HD state. Our model classified participants according to fine-grained (PreHD A; PreHD B; HD1; HD2), and binary disease states (PreHD vs HD; PreHD A vs PreHD B; PreHD B vs HD1; and HD1 vs HD2) to quantify feature importance across disease states.

Our main research questions were:

- Does our stacked ML model provide a systematic benefit for stratifying individuals according to HD states when using sMRI data?
- How many brain regions are required for optimal predictive classification, what are those regions, and do they vary with disease states?

We demonstrate that our stacked ML model is a powerful tool for early classification of fine-grained HD disease state with potential applications for clinical trial stratification, and can be easily extrapolated to other neurodegenerative diseases.

## Materials and methods

### Participants

We used baseline data from 184 participants from the TRACK-HD^3^ study (see Table 1). The data were collected at four different sites and included 104 PreHD and 80 manifest (HD).^3^ We excluded data from 16 PreHD and 43 HD participants as they failed visual QC.^48^ All imaging data were quality-controlled during conduct of the TRACK-HD study.

**Table 1:** Baseline cross-sectional demographic data for chosen subset of the TRACK-HD cohort.

| Demographics | Premanifest (PreHD) | Manifest (HD) |
| --- | --- | --- |
| N | 104 [58; 46] | 80 [49; 31] |
| Gender (0/1) | 55/49 | 43/37 |
| Age (years, mean ± SD) | 41.16±8.80 | 48.49±9.38 |
| CAG (repeats, mean ± SD) | 43.01±2.33 | 43.76±3.06 |

Participants were assigned into one of four classes as described in Tabrizi et al., 2009.^3^ Specifically, individuals without clinical HD symptoms but carrying the mutant HD gene were classed as PreHD A if they were >= 10.8 years from predicted onset, or were classed as PreHD B if they were < 10.8 years from predicted onset. Participants were classed as HD1 if they were diagnosed with clinical HD symptoms and had total functional capacity (TFC) score of 11-13, or were classed as HD2 if they had a TFC score of 7-10. There were n=58 participants in PreHD A, n=46 in PreHD B, n=49 HD1, and n=31 in HD2. These four groups cover a wide range of the HD progression spectrum and constitute a fine-grained assignment that distinguishes HD states both pre- and post-clinical manifestation of the symptoms.

These group classifications are derived from the original TRACK-HD study, which was conducted years before the novel Huntington disease integrated staging system (HD-ISS)^35^ became available. Therefore, the grouping and terminology does not follow the HD-ISS recommendations.

### Magnetic resonance imaging

Structural 3T T1-weighted MRI data were processed with The Geodesic Information Flows (GIF)^23^ software to segment and parcellate cortical and subcortical volumes. To avoid the influence of confounding factors that may set the groups apart, we corrected the volumes for age, research site, and intracranial volume (ICV) using linear regression.

### sMRI features

We used 15 sMRI volumetric measures that cover most of the cortical and subcortical regions. These included: *caudate, putamen, pallidum, thalamus, occipital lobe, lateral ventricles, frontal, temporal, accumbens area, insula, insula white matter, sensory motor, cingulate, parietal, and whole-brain*. The imaging features were ICV corrected and standard scaled.

There is a high degree of multicollinearity amongst these imaging features. Multicollinearity might not affect the accuracy of predictive models but poses a problem in the interpretability of the model; i.e., there is less reliability when determining the effects of specific features (independent predictors) on the dependent feature (targets).^24^ Despite this issue, we included all 15 sMRI features as we hypothesized that different sMRI features are differentially sensitive to HD-related brain alterations at different states; these brain alterations might progress at different rates or might interact with other structures differently in a multivariate context to mark progression to the next disease state.^40^

### Stacked ensemble ML-model

Ensemble models in ML combine decisions from multiple models to improve the overall performance.^25^ Research in ensemble methods field shows that they are more robust, reliable and accurate than standalone ML models. Single learners that conduct local searches may get stuck in local optima. By combining several learners, ensemble methods decrease the risk of obtaining a local minimum.^26^

Here, we use a powerful ensemble technique called the stacked ensemble model.^22^ The stacked model consists of two or more base models, also known as level 0 models and trainable meta-model. The base models were trained and evaluated using (repeated) k-fold cross-validation on actual training dataset. The predictions made by base models on out-of-sample data were then used to train the meta-model. The meta-model learns how to best combine the predictions made by the base models with the intent of reducing variance and generalisation error. The predictions made by meta-model are the final stacked model predictions. The schematic of stacked ensemble model is shown in supplementary material section S2.

Ensemble learning has been proven to produce improved and more robust performance than single models, and ergo its use is being increasingly explored in several neurodegenerative diseases such as Parkinson’s,^41–43^ Azheimers,^44, 45^ and multiple sclerosis.^46, 47^ However, to our knowledge, no other study has explored the use of stacked model for Huntington’s disease.

We designed a 2-tier stacked model, consisting of six standard ML models as base models – logistic regression (LR),^27^ K-nearest neighbours (KNN);^28^ support vector machine classifier (SVM);^29^ Gaussian naïve bayes (Bayes);^30^ Decision tree (cart);^31^ and multi-layer perceptron (MLP).^32^ We chose these models since these they are heterogeneous with varying strengths and characteristics; and ensured a good mixture of simple linear models such as LR and non-linear algorithms such as KNN, bayes, SVM, cart and MLP. We used a separate Gaussian naïve bayes model as the trainable meta-model.

All six base-models were trained and evaluated on the training dataset using repeated k-fold cross-validation. Their predictions on out-of-sample data were then combined using 5-fold cross-validation to create the training set for the meta-model. We did not perform any classifier-selection or hyperparameter tuning; the default parameter setting is described in supplementary material section S3.

### Cross-validation based model evaluation

To test the stability of the results and obtain robust estimates of prediction performance, we used repeated *stratified* k-fold cross-validation. Stratified sampling makes sure that class distribution in each split (i.e., fold) of data matches the distribution in the complete training dataset. Further, we *repeated* model training and testing multiple times with different 5-fold splits and report the average results from all repetitions. These repetitions mitigate extreme findings of high or low accuracy due to a single k-fold split, and ultimately produce more accurate, stable estimates of accuracy. Additionally, to handle class-imbalance within groups we employed ‘*balanced accuracy*’ to measure model performance. More details are in supplementary material section S4.

Additionally, we used dependent t-test for paired samples (ttest_rel function from python sklearn) to check significance of our results. We compared mean accuracy obtained for each repeat for stacked model with each of the base (& other) models i.e., 50 values for stacked model vs 50 values for each of the other model. We also correct p-values for multiple comparisons using Bonferroni correction throughout the paper.

### Quantifying feature importance

We performed feature importance analysis outside main striatal regions using a large number (15) of sMRI features We ranked brain regions by their importance for two reasons: 1) to aid the interpretation of the key regions that can be used as endpoints in future studies, and 2) to increase the accuracy of classification by removing unnecessary brain regions. We followed a greedy search approach using backward elimination. Specifically, we built a model with all the regions, then removed each region individually and computed the accuracy without that region, and then selected for removal of the region that caused the maximal increase (or the least decrease) in accuracy. This procedure was repeated recursively while removing one by one all but one last region.

This method provides the ordering of feature elimination and hence quantifies the relative importance of each feature. Details about this method are in the supplementary material section S5.

What differentiates our method from other HD classification studies (in supplementary material section S1) that employ feature selection a priori is that our method performs feature importance analysis via the classification tasks i.e., HD classification tasks are used to identify important features instead of the other way round which most studies do.

### Analysis design

#### Evaluation of the stacked model for HD progression classification

These analyses were conducted to explore the utility of the stacked ensemble model in classifying subjects into fine-grained HD states. The aim was to examine the suitability of a stacked ensemble approach to HD state classification and consequently assessing whether using stacked ensemble might result in an improvement on the current state-of-the-art for patient stratification. Instead of seeking an optimal (aka highest possible) classification accuracy for a given task as is typically the case, we aimed to conduct an exploratory analysis investigating potential benefits of stacked modelling approach on predictive accuracy and feature importance.

A total of 5 different classification tasks were performed, covering all HD states – premanifest vs manifest (PreHD vs HD); premanifest (PreHD A vs PreHD B); premanifest-manifest (PreHD B vs HD1); manifest (HD1 vs HD2); and finally fine-grained (PreHD A; PreHD B; HD1; HD2).

#### Identifying the brain regions with the best combined predictive accuracy

Next, for each classification task, we quantified feature importance to identify brain regions that carry the most discriminative information. The aim here was to investigate how many brain-regions are required for optimal predictive-classification of HD state, and what are those regions. We varied the number of repeats to evaluate the stacked model in terms of self-consistency and stability. This approach allowed us to investigate if the ordering of feature elimination is consistent across repetitions and, therefore, reliable. Even if the exact order of feature elimination is not the same across repeats, we hypothesized that the general trends should be similar. For each task, we report the (a) ordering in which features were eliminated in a given repeat; and (b) the classification accuracy and the standard deviation values at each feature elimination step (Tables 4.1 to 4.5).

## Data availability

Requests to access TRACK-HD dataset used in this study can be made to CHDI foundation: https://chdifoundation.org/policies/.

## Results

### Evaluation of the stacked model for HD progression classification

The experiment settings are described in table 2. We compared stacked model with its constituent base-models. To provide a more complete perspective, we also trained and tested three other popular ensemble models: Adaboost (Ada),^26^ random forest (RF),^26^ and extreme gradient boosting (XGB)^26^ using exactly the same data, cross-validation splits, and accuracy metrics.

**Table 2:**
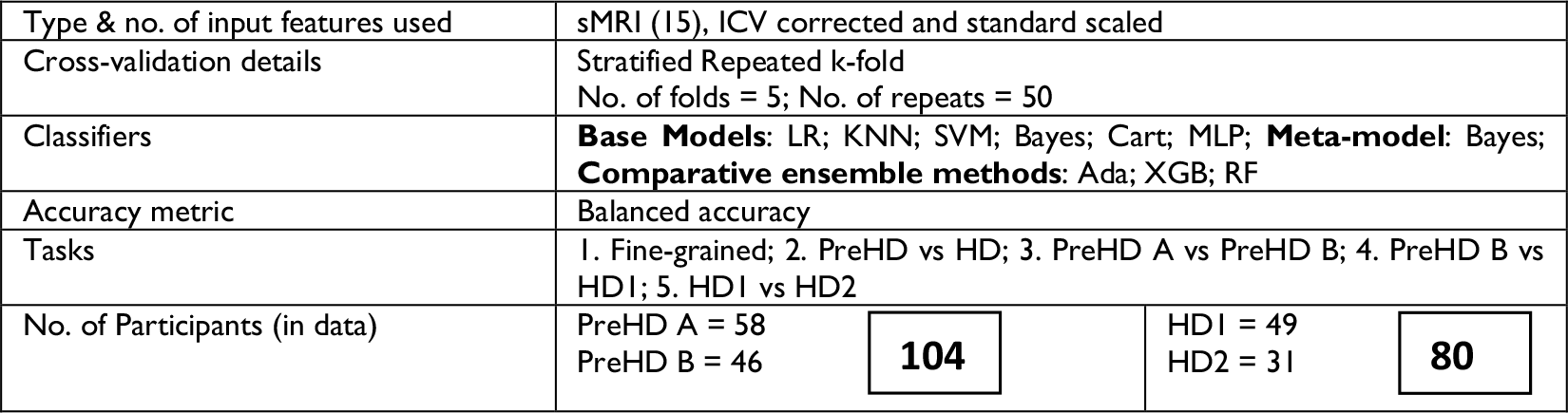
Experiment settings.

#### Premanifest-manifest (PreHD vs HD)

The base models distinguished PreHD from HD participants with mean accuracy varying between 72.3%±7.2 and 81.8%±6.2, whilst the stacked model attained 81.8%±5.9 accuracy. The stacked model performed as good as the best base models; however, it performs significantly better than most other models (*p<0.05* for knn, cart, Ada, RF, and XGB).

#### Premanifest (PreHD A vs PreHD B)

The base models correctly classified subjects into PreHD A and PreHD B classes with accuracies varying between 56.9%±10.6 and 68.6%±7.4. The stacked model achieved an accuracy of 68.6%±8.1, and performed significantly better than all but one base-models (i.e., Bayesian) and other comparative ensemble methods (*p <0.05*).

#### Premanifest-manifest (PreHD B vs HD1)

The base models classified subjects with accuracy between 60.9%±11.4 and 71.0%±9.0. The stacked model performance, 70.6%±9.7, was significantly better than all other models except logistic regression (*p<0.05*).

#### Manifest (HD1 vs HD2)

The same trend was observed for this task wherein the stacked model achieved an accuracy of 59.8%±10.8 whilst the base models’ accuracy varied between 52.2%±10.5 and 60.3%±11.7. The stacked model performed significantly better (*p<0.05*) than all models except LR.

#### Fine-grained (PreHD A; PreHD B; HD1; HD2)

The base models classified each participant as per their fine-grained disease state with mean accuracies between 40.2%±7.9 and 50.2%±5.6. In comparison, the stacked model achieved an accuracy of 52.6%±5.6, performing significantly better (*p<0.0005*) than all the other models. Performance details for all tasks such as accuracy values, standard deviations and p-values are included in supplementary material section S6.

Figure 1.1-1.5 compares the distribution of mean accuracy scores per repeat (i.e., 50 values per model) for each model, the base models are highlighted in pink, the stacked model is in green whereas the comparative ensemble approaches are in blue colour. In these plots, the box extends from the Q1 to Q3 quartile values of the data, with an orange line at the median (Q2). The whiskers extend from the edges of box to show the range of the data. By default, they extend no more than *1.5 * IQR (IQR = Q3 – Q1)* from the edges of the box, ending at the farthest data point within that interval. Outliers are plotted as separate dots. The green triangle represents the mean.

**Figure 1.1:**
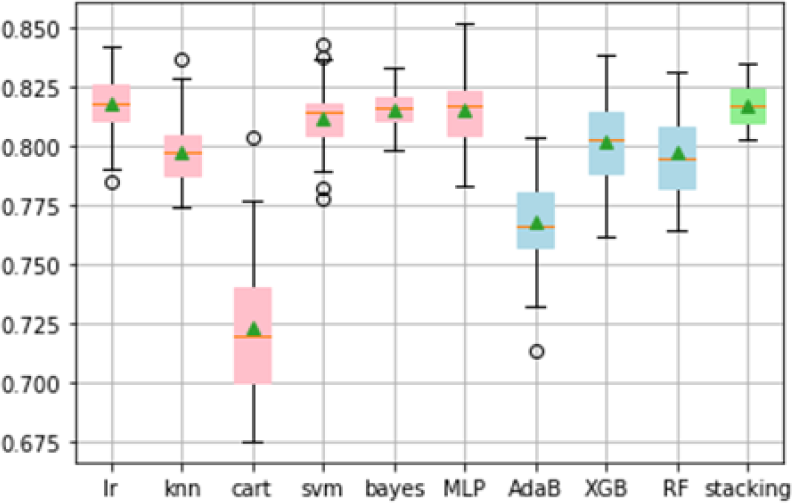
Accuracy distribution fo PreHD vs HD classification task.

**Figure 1.2:**
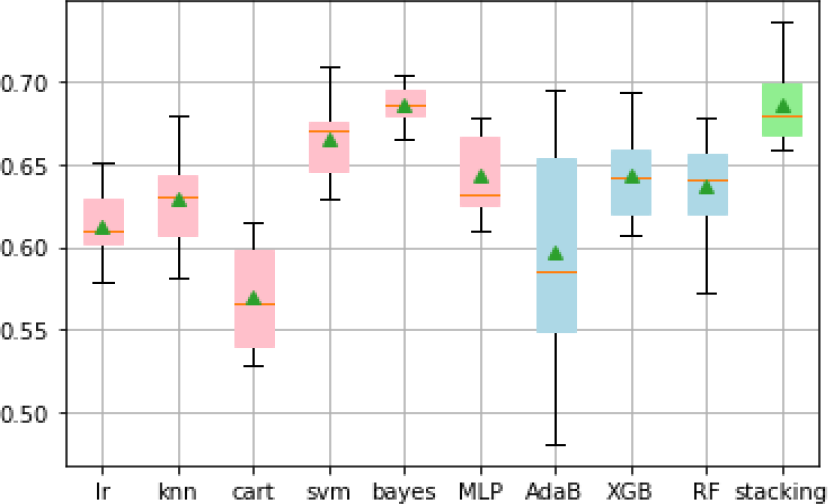
Accuracy scores distribution for PreHD A vs PreHD B.

**Figure 1.3:**
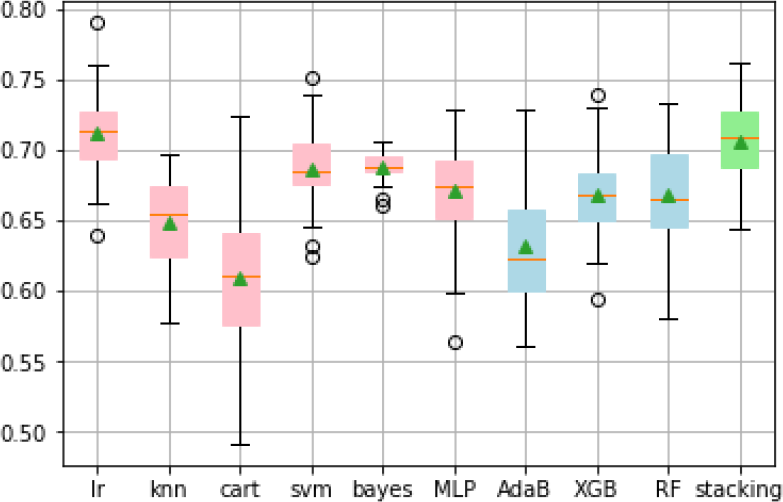
Accuracy scores distribution for PreHD B vs HD1 task.

**Figure 1.4:**
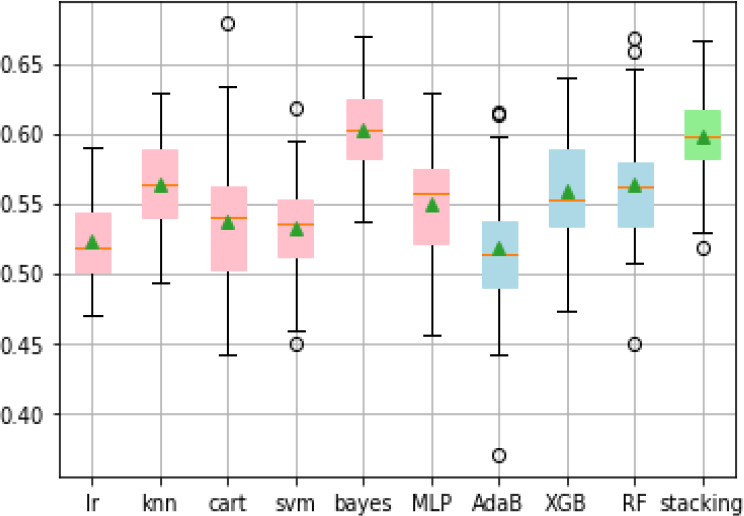
Accuracy scores distribution for HD1 vs HD2 task.

**Figure 1.5:**
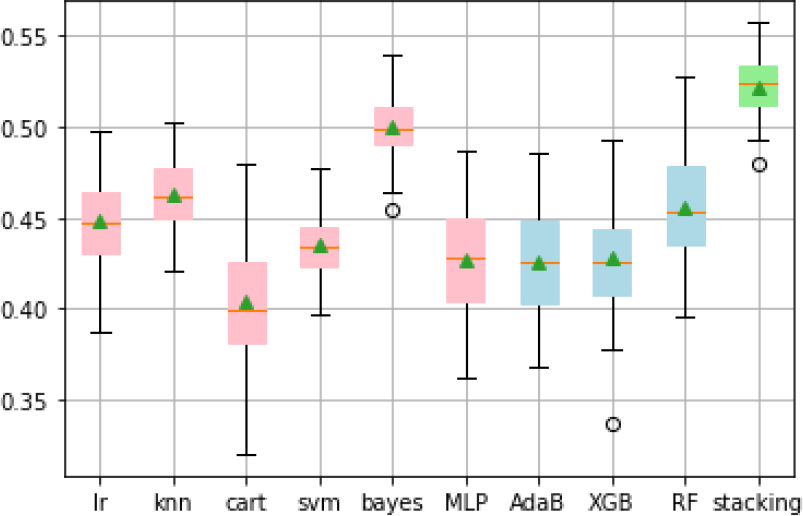
Accuracy scores distribution for fine-grained task.

We observed that stacked model has the most consistent performance and performed on average better than other models. Next, we aimed to identify the set of relevant brain regions for each task.

### Identifying the brain regions with the best combined predictive accuracy

The experiment settings are same as those described in Table 2, albeit with varying number of cross-validation repeats and hierarchical clustering of sMRI features described in Table 3.

**Table 3:** Experiment settings.

|  |  |
| --- | --- |
| Cross-validation details | Stratified Repeated k-fold<br>No. of folds = 5; No. of repeats = 10; 20; 30; 40; & 50 [~ 150] |
| Hierarchical clustering | Sklearn cluster.FeatureAgglomeration<br>Distance metric = Euclidean; linkage metric = ward |

In Tables 4.1 to 4.5, for a given no. of repeats, the columns represent the ordering of feature elimination starting with all features (n=15 in extreme left) to the single most important feature (n=1 in extreme right). The table cells display the mean accuracy and standard deviation (SD) attained by the stacked model at each step. For example, in Table 4.1 PreHD vs HD classification task, the model attained 80.4%±6.6 accuracy using all 15 features and eliminating Insula white matter resulted in maximum stacked model accuracy and hence it was discarded in that instance.

**Table 4.1:** Stacked model mean accuracy and standard deviation on PreHD vs HD classification.

|  |  |  |  |  |  |  |  |  |  |  |  |  |  |  |
| --- | --- | --- | --- | --- | --- | --- | --- | --- | --- | --- | --- | --- | --- | --- |
| **No. of repeats = 10** |  |  |  |  |  |  |  |  |  |  |  |  |  |  |
| 15 features | 14 | 13 | 12 | 11 | 10 | 9 | 8 | 7 | 6 | 5 | 4 | 3 | 2 | 1 |
| Insula WM | Insula | Wh. Brain | Pallidum | Thalamus | Temporal | Accumbens | SensMot. | Parietal | Putamen | Occipital | Cingulate | Frontal | Lat. Vents | Caudate |
| 0.804 | 0.819 | 0.817 | 0.822 | **0.832** | **0.832** | 0.828 | **0.833** | 0.825 | 0.818 | 0.822 | 0.821 | 0.825 | 0.807 | 0.795 |
| 0.066 | 0.053 | 0.059 | 0.062 | 0.052 | 0.060 | 0.056 | 0.047 | 0.056 | 0.060 | 0.049 | 0.051 | 0.045 | 0.061 | 0.064 |
| **No. of repeats = 20** |  |  |  |  |  |  |  |  |  |  |  |  |  |  |
| 15 features | 14 | 13 | 12 | 11 | 10 | 9 | 8 | 7 | 6 | 5 | 4 | 3 | 2 | 1 |
| Insula | Wh. Brain | Pallidum | Insula WM | Occipital | Frontal | Putamen | Thalamus | Parietal | Accumbens | Cingulate | Temporal | Lat. Vents | SensMot. | Caudate |
| 0.806 | 0.816 | 0.819 | 0.828 | **0.833** | 0.829 | 0.823 | 0.826 | 0.828 | 0.825 | 0.812 | 0.819 | 0.815 | 0.812 | 0.800 |
| 0.064 | 0.056 | 0.057 | 0.054 | 0.063 | 0.062 | 0.060 | 0.061 | 0.063 | 0.067 | 0.061 | 0.065 | 0.057 | 0.060 | 0.060 |
| **No. of repeats = 30** |  |  |  |  |  |  |  |  |  |  |  |  |  |  |
| 15 features | 14 | 13 | 12 | 11 | 10 | 9 | 8 | 7 | 6 | 5 | 4 | 3 | 2 | 1 |
| Insula | Insula WM | Occipital | Frontal | Wh. Brain | Pallidum | Putamen | Thalamus | Parietal | Accumbens | Lat. Vents | Cingulate | SensMot. | Temporal | Caudate |
| 0.809 | 0.816 | 0.819 | 0.824 | 0.828 | **0.831** | 0.823 | 0.823 | **0.832** | 0.828 | 0.817 | 0.818 | 0.818 | 0.817 | 0.799 |
| 0.055 | 0.056 | 0.058 | 0.052 | 0.059 | 0.059 | 0.054 | 0.060 | 0.052 | 0.060 | 0.054 | 0.062 | 0.064 | 0.059 | 0.062 |
| **No. of repeats = 40** |  |  |  |  |  |  |  |  |  |  |  |  |  |  |
| 15 features | 14 | 13 | 12 | 11 | 10 | 9 | 8 | 7 | 6 | 5 | 4 | 3 | 2 | 1 |
| Insula | Insula WM | Wh. Brain | Pallidum | Occipital | Thalamus | Parietal | Frontal | Putamen | Accumbens | Lat. Vents | Cingulate | SensMot. | Temporal | Caudate |
| 0.806 | 0.814 | 0.817 | 0.821 | **0.829** | 0.827 | 0.827 | **0.829** | **0.829** | **0.828** | 0.815 | 0.818 | 0.817 | 0.817 | 0.796 |
| 0.062 | 0.057 | 0.058 | 0.062 | 0.055 | 0.053 | 0.056 | 0.056 | 0.051 | 0.060 | 0.060 | 0.054 | 0.062 | 0.055 | 0.060 |
| **No. of repeats = 50** |  |  |  |  |  |  |  |  |  |  |  |  |  |  |
| 15 features | 14 | 13 | 12 | 11 | 10 | 9 | 8 | 7 | 6 | 5 | 4 | 3 | 2 | 1 |
| Insula | Occipital | Pallidum | Wh. Brain | Insula WM | Thalamus | Parietal | Frontal | Putamen | Accumbens | Lat. Vents | Cingulate | SensMot. | Temporal | Caudate |
| 0.806 | 0.816 | 0.818 | 0.823 | 0.827 | 0.828 | 0.827 | **0.830** | **0.830** | 0.824 | 0.817 | 0.820 | 0.817 | 0.821 | 0.797 |
| 0.063 | 0.059 | 0.059 | 0.062 | 0.056 | 0.058 | 0.056 | 0.054 | 0.056 | 0.053 | 0.062 | 0.061 | 0.054 | 0.059 | 0.057 |

**Table 4.2:**
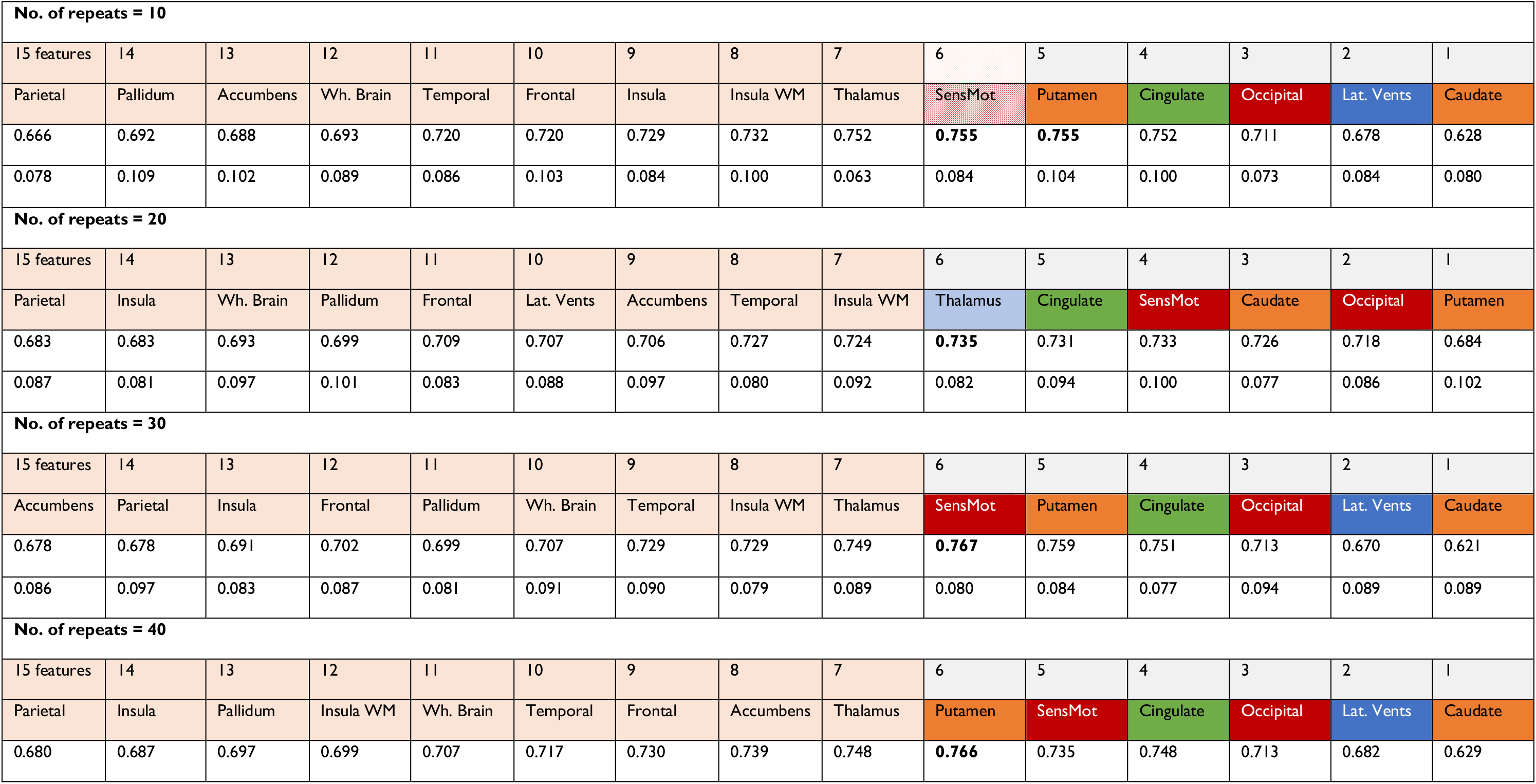
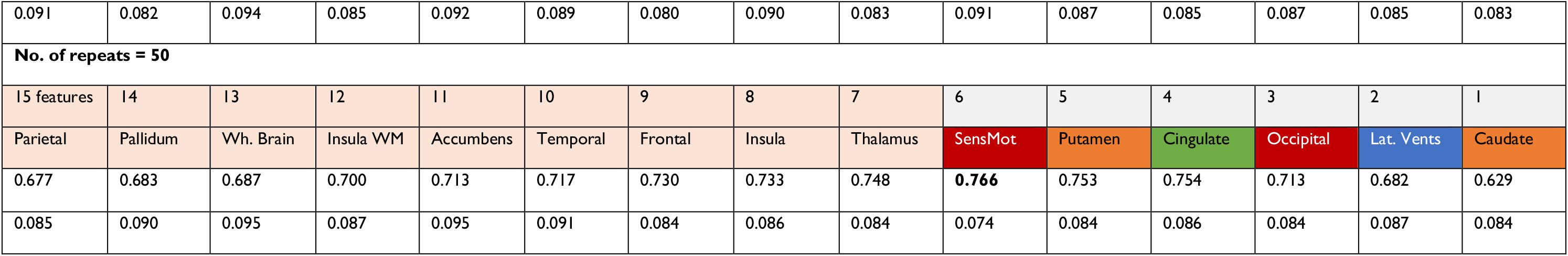
Stacked model mean accuracy & standard deviation on PreHD A vs PreHD B task.

**Table 4.3:** Stacked model mean accuracy & standard deviation on PreHD B vs HD1 task.

|  |  |  |  |  |  |  |  |  |  |  |  |  |  |  |
| --- | --- | --- | --- | --- | --- | --- | --- | --- | --- | --- | --- | --- | --- | --- |
| **No. of repeats = 10** |  |  |  |  |  |  |  |  |  |  |  |  |  |  |
| 15 features | 14 | 13 | 12 | 11 | 10 | 9 | 8 | 7 | 6 | 5 | 4 | 3 | 2 | 1 |
| Insula | Pallidum | Insula WM | Thalamus | Occipital | Putamen | Wh. Brain | Parietal | SensMot | Accumbens | Lat. Vents | Frontal | Cingulate | Temporal | Caudate |
| 0.688 | 0.693 | 0.698 | 0.684 | 0.685 | 0.702 | 0.679 | 0.703 | 0.702 | **0.742** | 0.725 | 0.708 | 0.726 | 0.707 | 0.688 |
| 0.101 | 0.101 | 0.100 | 0.105 | 0.117 | 0.094 | 0.108 | 0.097 | 0.093 | 0.094 | 0.075 | 0.111 | 0.089 | 0.077 | 0.102 |
| **No. of repeats = 20** |  |  |  |  |  |  |  |  |  |  |  |  |  |  |
| 15 features | 14 | 13 | 12 | 11 | 10 | 9 | 8 | 7 | 6 | 5 | 4 | 3 | 2 | 1 |
| Insula | Thalamus | Pallidum | Accumbens | Occipital | Wh. Brain | SensMot | Putamen | Parietal | Insula WM | Lat. Vents | Frontal | Cingulate | Temporal | Caudate |
| 0.693 | 0.700 | 0.690 | 0.701 | 0.701 | 0.707 | 0.719 | 0.722 | 0.718 | 0.731 | **0.735** | 0.709 | 0.710 | 0.705 | 0.691 |
| 0.090 | 0.088 | 0.090 | 0.106 | 0.107 | 0.096 | 0.088 | 0.087 | 0.094 | 0.097 | 0.092 | 0.104 | 0.088 | 0.093 | 0.096 |
| **No. of repeats = 30** |  |  |  |  |  |  |  |  |  |  |  |  |  |  |
| 15 features | 14 | 13 | 12 | 11 | 10 | 9 | 8 | 7 | 6 | 5 | 4 | 3 | 2 | 1 |
| Insula | Insula WM | Accumbens | Pallidum | Thalamus | SensMot | Parietal | Occipital | Putamen | Wh. Brain | Cingulate | Temporal | Lat. Vents | Frontal | Caudate |
| 0.690 | 0.696 | 0.688 | 0.694 | 0.698 | 0.691 | 0.695 | 0.713 | 0.705 | 0.706 | **0.739** | 0.719 | 0.716 | 0.705 | 0.691 |
| 0.092 | 0.100 | 0.098 | 0.092 | 0.090 | 0.084 | 0.098 | 0.094 | 0.105 | 0.092 | 0.089 | 0.083 | 0.099 | 0.087 | 0.089 |

| No. of repeats = 40 |  |  |  |  |  |  |  |  |  |  |  |  |  |  |
| --- | --- | --- | --- | --- | --- | --- | --- | --- | --- | --- | --- | --- | --- | --- |
| 15 features | 14 | 13 | 12 | 11 | 10 | 9 | 8 | 7 | 6 | 5 | 4 | 3 | 2 | 1 |
| Wh. Brain | Pallidum | Insula | Lat. Vents | Parietal | SensMot | Temporal | Putamen | Insula WM | Thalamus | Frontal | Accumbens | Cingulate | Occipital | Caudate |
| 0.690 | 0.696 | 0.700 | 0.704 | 0.714 | 0.717 | 0.726 | 0.719 | 0.727 | 0.712 | 0.712 | 0.711 | 0.711 | 0.697 | 0.691 |
| 0.095 | 0.090 | 0.094 | 0.088 | 0.098 | 0.100 | 0.090 | 0.082 | 0.095 | 0.097 | 0.099 | 0.096 | 0.089 | 0.102 | 0.089 |
| No. of repeats = 50 |  |  |  |  |  |  |  |  |  |  |  |  |  |  |
| 15 features | 14 | 13 | 12 | 11 | 10 | 9 | 8 | 7 | 6 | 5 | 4 | 3 | 2 | 1 |
| Putamen | Accumbens | Insula WM | Pallidum | Occipital | Thalamus | Parietal | Lat. Vents | Wh. Brain | Frontal | Cingulate | SensMot | Insula | Temporal | Caudate |
| 0.691 | 0.700 | 0.701 | 0.714 | 0.716 | 0.729 | 0.731 | 0.751 | 0.759 | 0.756 | 0.737 | 0.731 | 0.749 | 0.712 | 0.685 |
| 0.094 | 0.088 | 0.094 | 0.088 | 0.097 | 0.087 | 0.098 | 0.094 | 0.085 | 0.091 | 0.090 | 0.095 | 0.090 | 0.084 | 0.094 |

**Table 4.4:** Stacked model mean accuracy & standard deviation on HD1 vs HD2 task.

| No. of repeats = 10 |  |  |  |  |  |  |  |  |  |  |  |  |  |  |
| --- | --- | --- | --- | --- | --- | --- | --- | --- | --- | --- | --- | --- | --- | --- |
| 15 features | 14 | 13 | 12 | 11 | 10 | 9 | 8 | 7 | 6 | 5 | 4 | 3 | 2 | 1 |
| Insula WM | Pallidum | Accumbens | Putamen | Temporal | Cingulate | Wh. Brain | Frontal | Parietal | SensMot | Lat. Vents | Thalamus | Insula | Occipital | Caudate |
| 0.613 | 0.584 | 0.632 | 0.616 | 0.627 | 0.635 | 0.632 | 0.651 | 0.641 | 0.641 | 0.619 | 0.622 | 0.657 | **0.672** | 0.619 |
| 0.122 | 0.092 | 0.110 | 0.089 | 0.095 | 0.105 | 0.080 | 0.110 | 0.114 | 0.119 | 0.119 | 0.091 | 0.080 | 0.097 | 0.074 |
| No. of repeats = 20 |  |  |  |  |  |  |  |  |  |  |  |  |  |  |
| 15 features | 14 | 13 | 12 | 11 | 10 | 9 | 8 | 7 | 6 | 5 | 4 | 3 | 2 | 1 |
| Thalamus | Lat. Vents | Insula | Pallidum | Insula WM | Cingulate | Putamen | Accumbens | Wh. Brain | Frontal | Parietal | SensMot | Temporal | Occipital | Caudate |
| 0.592 | 0.592 | 0.623 | 0.641 | 0.632 | 0.641 | 0.645 | 0.660 | 0.674 | **0.683** | **0.682** | 0.672 | **0.680** | 0.673 | 0.624 |
| 0.109 | 0.120 | 0.114 | 0.110 | 0.103 | 0.103 | 0.104 | 0.108 | 0.102 | 0.108 | 0.108 | 0.103 | 0.121 | 0.102 | 0.100 |
| No. of repeats = 30 |  |  |  |  |  |  |  |  |  |  |  |  |  |  |
| 15 features | 14 | 13 | 12 | 11 | 10 | 9 | 8 | 7 | 6 | 5 | 4 | 3 | 2 | 1 |
| Insula WM | Pallidum | Accumbens | Wh. brain | Temporal | Cingulate | Putamen | Parietal | Frontal | SensMot | Insula | Lat. Vents | Thalamus | Occipital | Caudate |
| 0.589 | 0.615 | 0.633 | 0.642 | 0.631 | 0.645 | 0.660 | **0.679** | 0.655 | 0.671 | **0.675** | 0.648 | 0.659 | 0.648 | 0.638 |
| 0.115 | 0.109 | 0.106 | 0.101 | 0.114 | 0.112 | 0.112 | 0.118 | 0.110 | 0.104 | 0.111 | 0.116 | 0.115 | 0.111 | 0.110 |
| **No. of repeats = 40** |  |  |  |  |  |  |  |  |  |  |  |  |  |  |
| 15 features | 14 | 13 | 12 | 11 | 10 | 9 | 8 | 7 | 6 | 5 | 4 | 3 | 2 | 1 |
| Pallidum | Insula WM | Accumbens | Wh. Brain | Cingulate | Temporal | Frontal | Insula | Putamen | Lat. Vents | Thalamus | Occipital | SensMot | Parietal | Caudate |
| 0.599 | 0.601 | 0.634 | 0.634 | 0.639 | 0.651 | 0.663 | 0.686 | 0.655 | 0.670 | 0.669 | 0.668 | **0.699** | 0.697 | 0.587 |
| 0.100 | 0.113 | 0.111 | 0.115 | 0.113 | 0.112 | 0.115 | 0.100 | 0.115 | 0.104 | 0.112 | 0.116 | 0.098 | 0.104 | 0.105 |
| **No. of repeats = 50** |  |  |  |  |  |  |  |  |  |  |  |  |  |  |
| 15 features | 14 | 13 | 12 | 11 | 10 | 9 | 8 | 7 | 6 | 5 | 4 | 3 | 2 | 1 |
| Insula WM | Pallidum | Accumbens | Wh. Brain | Temporal | Cingulate | Frontal | Putamen | Parietal | SensMot | Insula | Lat. Vents | Thalamus | Occipital | Caudate |
| 0.598 | 0.621 | 0.628 | 0.644 | 0.641 | 0.644 | 0.666 | 0.674 | 0.670 | 0.677 | 0.668 | 0.670 | 0.702 | **0.709** | 0.603 |
| 0.103 | 0.109 | 0.111 | 0.115 | 0.113 | 0.109 | 0.110 | 0.105 | 0.116 | 0.102 | 0.111 | 0.114 | 0.110 | 0.094 | 0.100 |

**Table 4.5:**
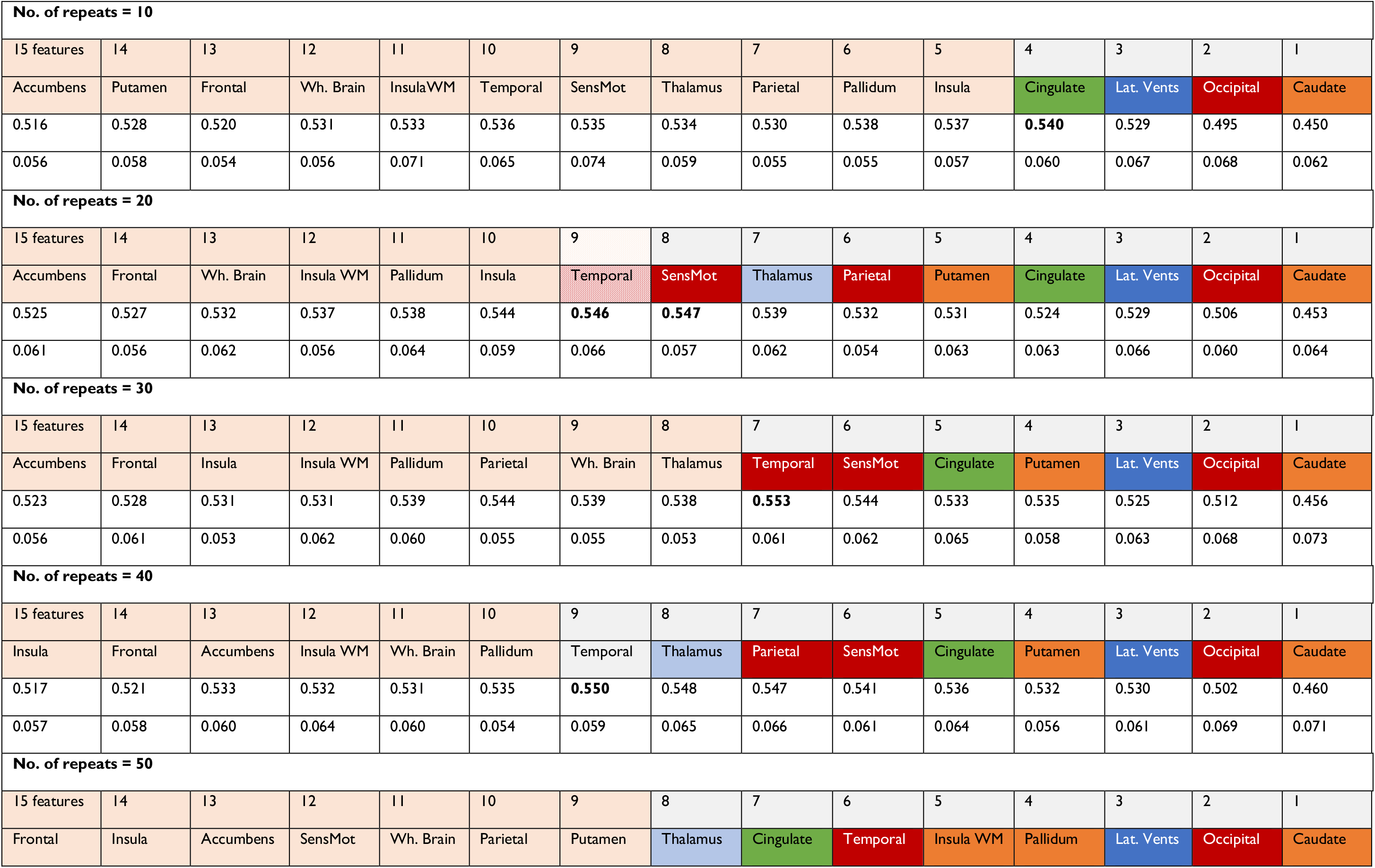
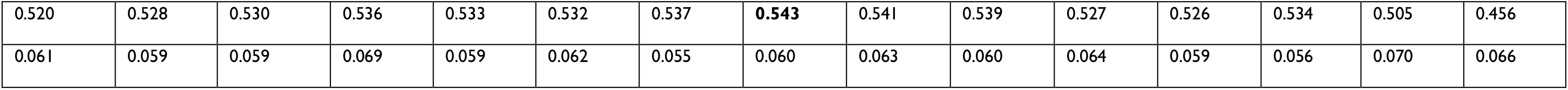
Stacked model mean accuracy and standard deviation on Fine-Grained task.

|  |  |  |  |  |  |  |  |  |  |  |  |  |  |  |
| --- | --- | --- | --- | --- | --- | --- | --- | --- | --- | --- | --- | --- | --- | --- |
| **No. of repeats = 10** |  |  |  |  |  |  |  |  |  |  |  |  |  |  |
| 15 features | 14 | 13 | 12 | 11 | 10 | 9 | 8 | 7 | 6 | 5 | 4 | 3 | 2 | 1 |
| Accumbens | Putamen | Frontal | Wh. Brain | InsulaWM | Temporal | SensMot | Thalamus | Parietal | Pallidum | Insula | Cingulate | Lat. Vents | Occipital | Caudate |
| 0.516 | 0.528 | 0.520 | 0.531 | 0.533 | 0.536 | 0.535 | 0.534 | 0.530 | 0.538 | 0.537 | **0.540** | 0.529 | 0.495 | 0.450 |
| 0.056 | 0.058 | 0.054 | 0.056 | 0.071 | 0.065 | 0.074 | 0.059 | 0.055 | 0.055 | 0.057 | 0.060 | 0.067 | 0.068 | 0.062 |
| **No. of repeats = 20** |  |  |  |  |  |  |  |  |  |  |  |  |  |  |
| 15 features | 14 | 13 | 12 | 11 | 10 | 9 | 8 | 7 | 6 | 5 | 4 | 3 | 2 | 1 |
| Accumbens | Frontal | Wh. Brain | Insula WM | Pallidum | Insula | Temporal | SensMot | Thalamus | Parietal | Putamen | Cingulate | Lat. Vents | Occipital | Caudate |
| 0.525 | 0.527 | 0.532 | 0.537 | 0.538 | 0.544 | **0.546** | **0.547** | 0.539 | 0.532 | 0.531 | 0.524 | 0.529 | 0.506 | 0.453 |
| 0.061 | 0.056 | 0.062 | 0.056 | 0.064 | 0.059 | 0.066 | 0.057 | 0.062 | 0.054 | 0.063 | 0.063 | 0.066 | 0.060 | 0.064 |
| **No. of repeats = 30** |  |  |  |  |  |  |  |  |  |  |  |  |  |  |
| 15 features | 14 | 13 | 12 | 11 | 10 | 9 | 8 | 7 | 6 | 5 | 4 | 3 | 2 | 1 |
| Accumbens | Frontal | Insula | Insula WM | Pallidum | Parietal | Wh. Brain | Thalamus | Temporal | SensMot | Cingulate | Putamen | Lat. Vents | Occipital | Caudate |
| 0.523 | 0.528 | 0.531 | 0.531 | 0.539 | 0.544 | 0.539 | 0.538 | **0.553** | 0.544 | 0.533 | 0.535 | 0.525 | 0.512 | 0.456 |
| 0.056 | 0.061 | 0.053 | 0.062 | 0.060 | 0.055 | 0.055 | 0.053 | 0.061 | 0.062 | 0.065 | 0.058 | 0.063 | 0.068 | 0.073 |
| **No. of repeats = 40** |  |  |  |  |  |  |  |  |  |  |  |  |  |  |
| 15 features | 14 | 13 | 12 | 11 | 10 | 9 | 8 | 7 | 6 | 5 | 4 | 3 | 2 | 1 |
| Insula | Frontal | Accumbens | Insula WM | Wh. Brain | Pallidum | Temporal | Thalamus | Parietal | SensMot | Cingulate | Putamen | Lat. Vents | Occipital | Caudate |
| 0.517 | 0.521 | 0.533 | 0.532 | 0.531 | 0.535 | **0.550** | 0.548 | 0.547 | 0.541 | 0.536 | 0.532 | 0.530 | 0.502 | 0.460 |
| 0.057 | 0.058 | 0.060 | 0.064 | 0.060 | 0.054 | 0.059 | 0.065 | 0.066 | 0.061 | 0.064 | 0.056 | 0.061 | 0.069 | 0.071 |
| **No. of repeats = 50** |  |  |  |  |  |  |  |  |  |  |  |  |  |  |
| 15 features | 14 | 13 | 12 | 11 | 10 | 9 | 8 | 7 | 6 | 5 | 4 | 3 | 2 | 1 |
| Frontal | Insula | Accumbens | SensMot | Wh. Brain | Parietal | Putamen | Thalamus | Cingulate | Temporal | Insula WM | Pallidum | Lat. Vents | Occipital | Caudate |
| 0.520 | 0.528 | 0.530 | 0.536 | 0.533 | 0.532 | 0.537 | **0.543** | 0.541 | 0.539 | 0.527 | 0.526 | 0.534 | 0.505 | 0.456 |
| 0.061 | 0.059 | 0.059 | 0.069 | 0.059 | 0.062 | 0.055 | 0.060 | 0.063 | 0.060 | 0.064 | 0.059 | 0.056 | 0.070 | 0.066 |

The features highlighted in pink are the ones whose elimination resulted in increased classification accuracy for that number of CV repeats. Highest accuracy was achieved when a certain number of input features were removed; that inflection point was the ‘highest accuracy value’, and consequently removing the subset of features in pink gave us the optimal set of features (highlighted in different bold ‘cluster’ colours) required by the stacked model to achieve maximal accuracy.

Tables 4.1-4.5 show that the removal of brain regions with backward elimination did not always follow the same order. We hypothesized that the instability is caused by correlated variables that flip their position, i.e., one correlated variable being removed earlier and the other later, or vice versa, depending on the splits into folds. To investigate this hypothesis, we ran hierarchical clustering and produced a dendrogram tree of all the variables via python sklearn’s built-in dendrogram and agglomerative function cluster.FeatureAgglomeration. The result is shown in Figure 2/Table 5.

**Figure 2:**
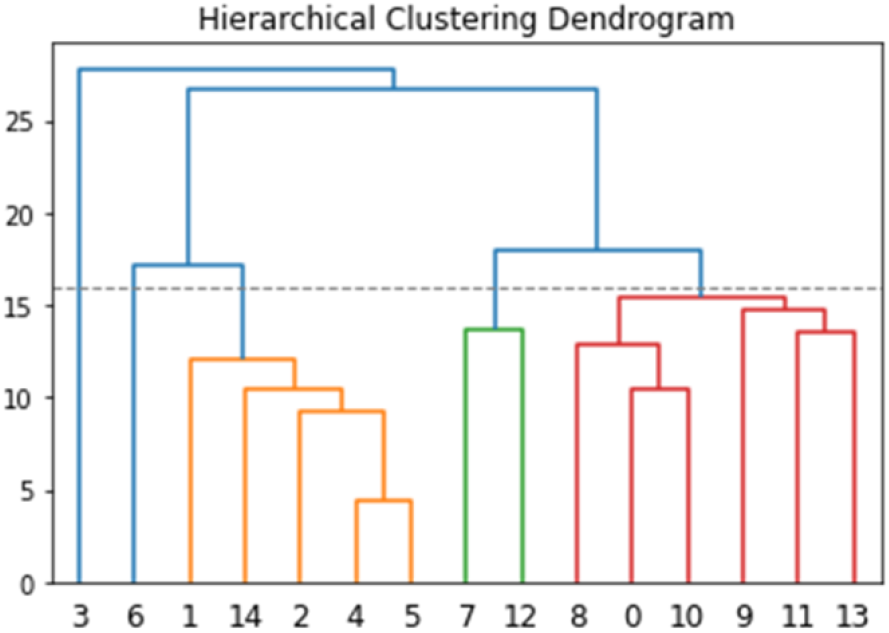
Dendrogram example.

**Table 5:**
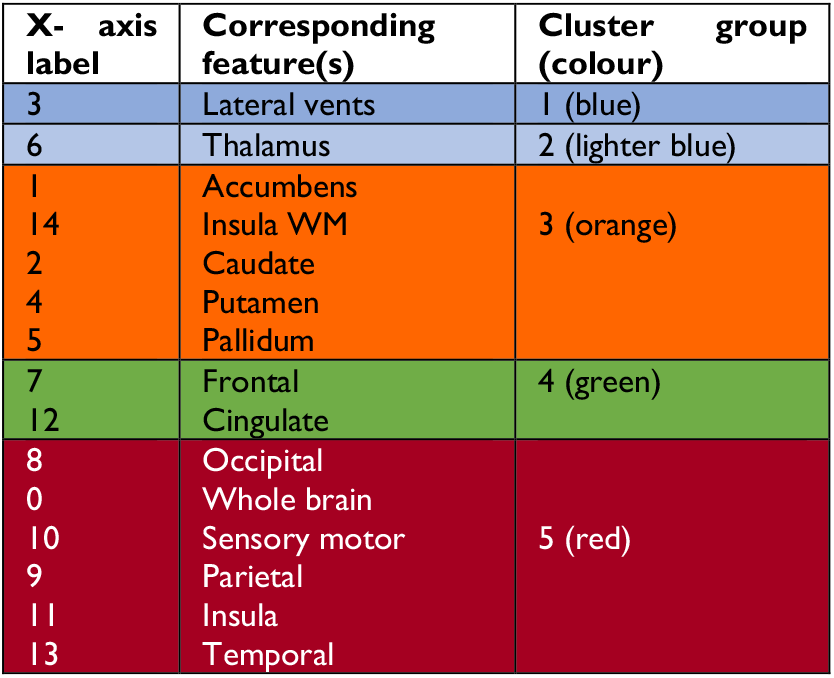
Feature agglomeration.

Further, one of our main goals was to identify which brain regions carry the most discriminative information for each disease state. We varied the number of repeats and applied the recursive feature elimination approach to determine if sequence of feature elimination is stable and consistent across repetitions. We hypothesized that, even though the exact order may not be the same, positional stability towards bottom right of heatmaps would be indicative of the most informative brain regions for that task.

Our results (in tables 4.1-4.5) showed that feature elimination sequences were not exactly the same in different repeats. Thus, to establish the consistency of the feature elimination order, we plotted positional heatmaps, figures 3.1-3.5. In each figure, dark diagonal component indicates more stable positioning at that position. Features were assigned a position according to the maximum likelihood sequence (i.e., baseline: y-axis), which represented the maximum probability of a feature getting eliminated at a particular position, i.e., y-axis ordering represents increasing feature importance from top-to-bottom. Details in supplementary material, section S7.

#### Premanifest-manifest classification (PreHD vs HD)

Table 4.1 and Figure 3.1 depict that our stacked model discriminated between PreHD and HD subjects with a maximum accuracy of 83%±6.3 and requires a range of 8-11 sMRI features to attain the highest accuracy. Caudate is consistently the most important feature across all repeats as it never gets eliminated before the maximal inflection point. Both the accuracy levels and the caudate winning the elimination procedure were stable and consistent across repetitions.

**Figure 3.1:**
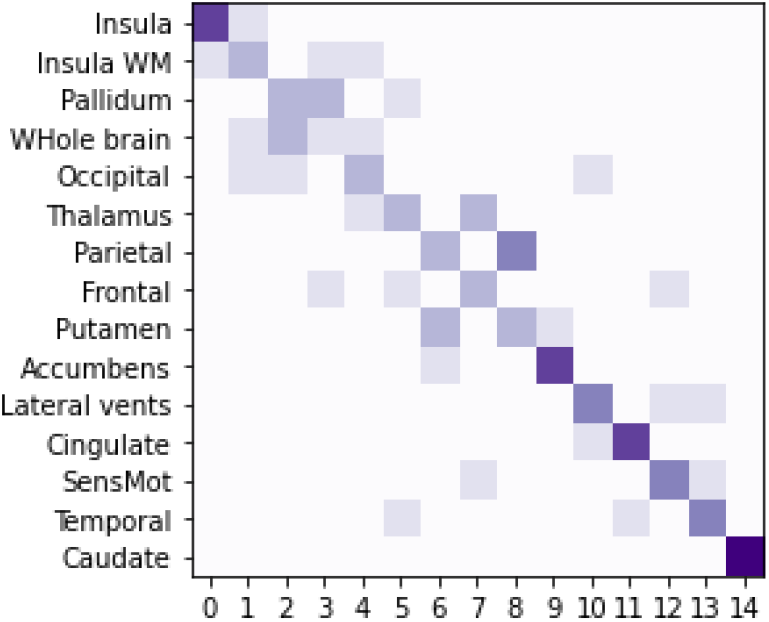
Feature elimination order across repeats PreHD vs HD task.

The maximum accuracy attained by the stacked model is not significantly different from accuracy with just caudate. Out of these 8-11 features, a subset of 6 brain regions (caudate, temporal, sensory motor, cingulate, lateral ventricles and accumbens) were consistently stable in their respective positions for >120 repeats (out of cumulative 150 repeats).

There was some positional variability (in < 30 repeats), but even in those cases, these features vary between near adjacent positions such as cingulate or lateral ventricles or they were still positioned among 8-11 informative brain regions (i.e., ‘cluster’ coloured cells in table 4.1), for example temporal, sensory motor and Accumbens area.

#### Premanifest (PreHD A vs PreHD B) classification

The classification accuracy using just the caudate was significantly lower than the highest accuracy of 76.7%±8.0, which required a set of 6 sMRI measures– caudate, occipital lobe, lateral ventricles, putamen, cingulate and sensory motor, all of which were positionally stable across repetitions (table 4.2).

Figure 3.2 highlights that variability in feature importance ordering is confined to *non-relevant* set of features i.e., top-half of diagonal; whilst for the set of important features it is stable and consistent across repeats and only varies a little between near adjacent positions only.

**Figure 3.2:**
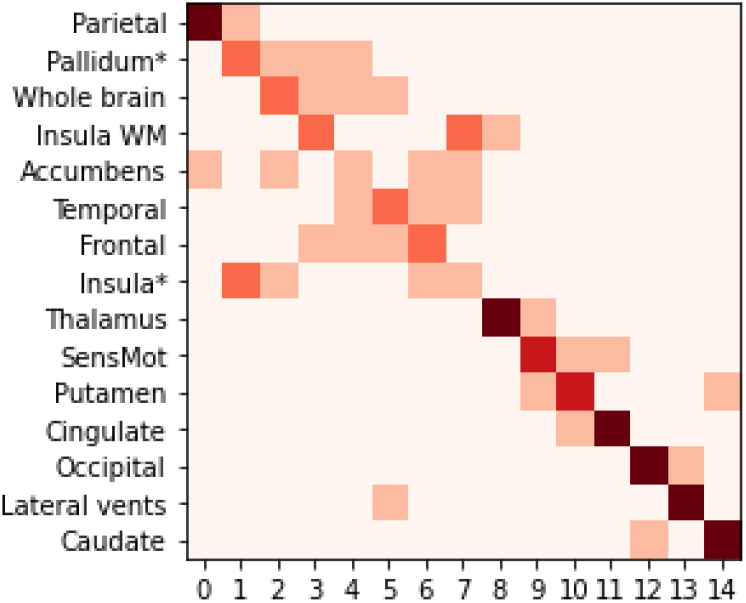
Feature elimination order across repeats PreHD A vs PreHD B task.

#### Premanifest–manifest (PreHD B vs HD1) classification

Table 4.3 depicts that stacked model segregates subjects into PreHD B and HD1 classes with an accuracy of approximately 69% by using caudate. The model attained the maximum accuracy of 75%±9.0 with at least 5-7 sMRI features, relying more on features belonging to cortical region and frontal-cingulate cluster. This result replicated across repetitions.

We found a high degree of uncertainty in feature elimination ordering for this classification task (figure 3.3). Only caudate and temporal are positionally stable for 100+ repeats.

**Figure 3.3:**
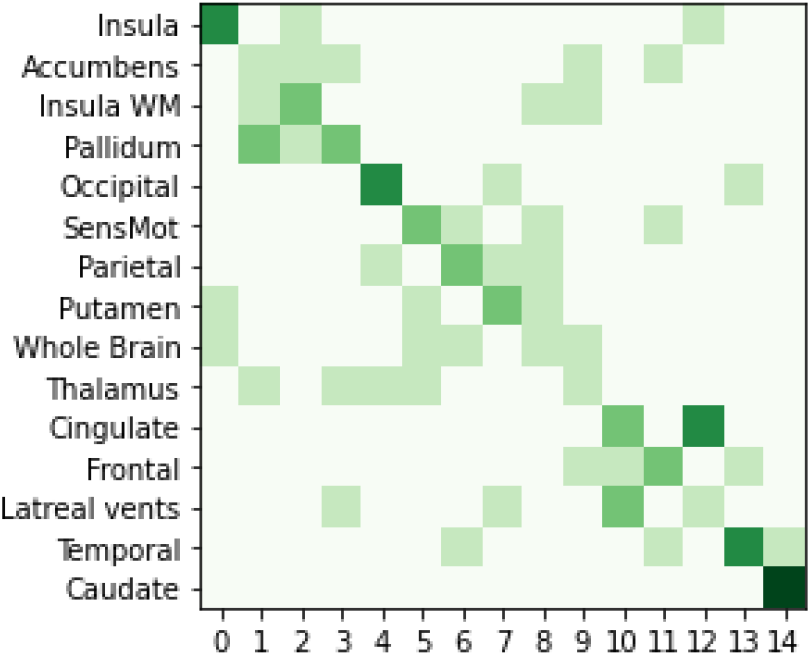
Feature elimination order across repeats PreHD B vs HD1.

#### Manifest (HD1 vs HD2) classification

The maximum accuracy of 68%±10.0 required between 2-5 features belonging to subcortical striatal and cortical regions, such as caudate and occipital lobe (Table 4.4). Further, figure 3.4 shows that the order of elimination is positionally stable towards the start (top-left) and end (bottom-right) wherein most features are placed strongly at diagonal. The more important features, i.e., those getting eliminated later (such as occipital, thalamus and lateral ventricles) vary within near-adjacent positions.

**Figure 3.4:**
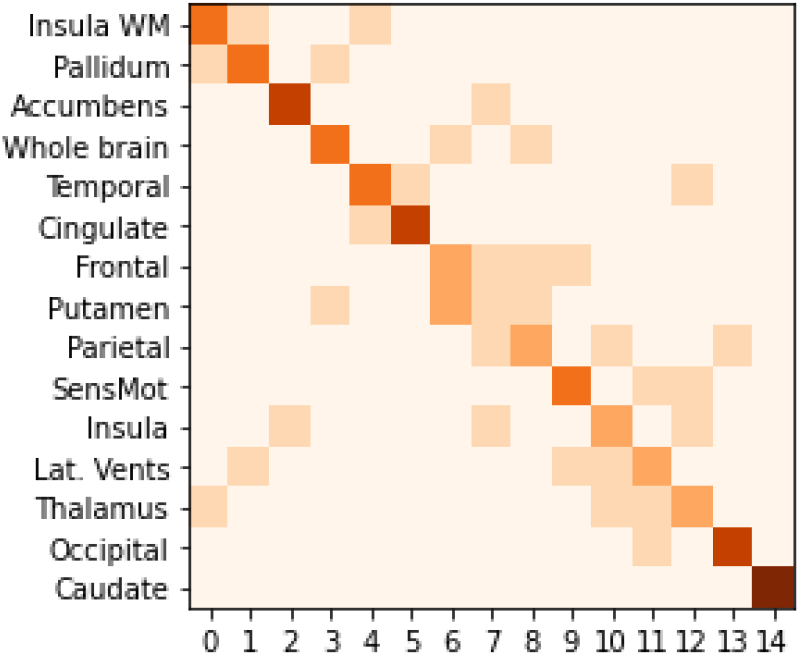
Feature elimination order across repeats HD1 vs HD2.

#### Fine-grained (PreHD A; PreHD B; HD1; HD2) classification

Our model achieved the maximum accuracy of 55%±6.0 and caudate, lateral ventricles and occipital lobe were certainly required to attain this accuracy across all repeats; albeit maximum accuracy required at least 2-5 other sMRI features.

Finally, it is worth noting that despite the instability, the set of more relevant features i.e., those contributing towards highest accuracy – occipital lobe, lateral ventricles and caudate remained stable across repetitions as seen in figure 3.5.

**Figure 3.5:**
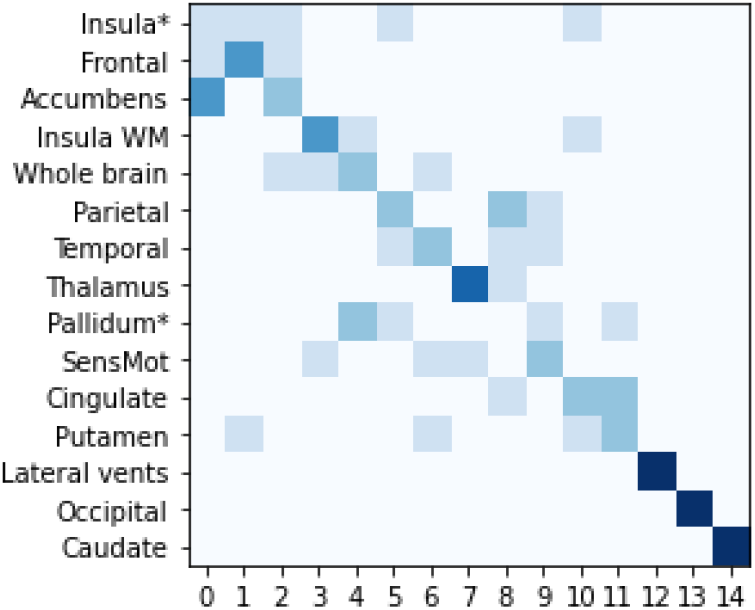
Feature elimination order across repeats Fine-grained task.

Our analyses show that there is instability in the exact order of feature elimination. However, the brain regions with the *best combined predictive accuracy* for each disease state remain the same across repetitions. Based on these results, we identified the brain regions with the best combined predictive accuracy, for each disease state, as the sMRI features that were amongst the last ones to get eliminated, i.e., bottom-right of heatmaps (or positioned in bold coloured cells in tables 4.1 to 4.5) in >= 100 repeats (i.e., more than 66.66%).

Each region’s contribution is vital to a feature subset informative for a particular classification task. These subsets of regions are specific for each task and were essential to attain highest predictive-classification accuracy. Some other features might be required in addition to these; however, those additional features vary with task and are unstable across repeats and therefore we did not include them as part of most informative brain regions. Table 6 makes it evident that: (1.) predictive-classification accuracy can be *improved* upon by including brain regions outside the striatum; and (2.) for optimal prediction stacked model tends to rely on sMRI features from different yet complementary feature clusters.

**Table 6:**
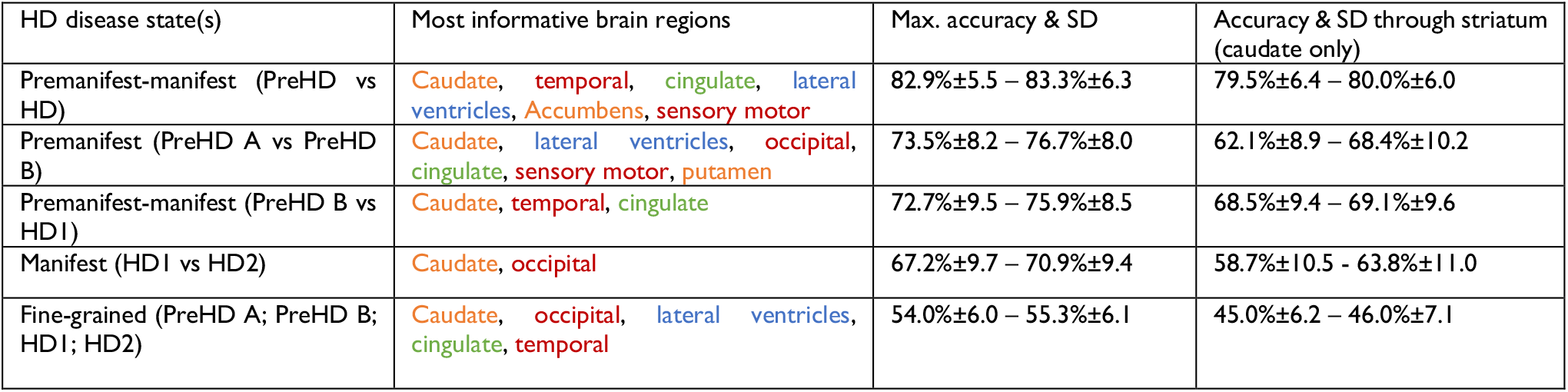
Set of brain regions with the best combined predictive accuracy for each HD disease state.

## Discussion

We investigated the utility of a stacked ensemble model for fine-grained classification of HD states using solely volumetric scores from sMRI. Overall, the results showed that more accurate predictions of disease progression state can be achieved by including structures outside the striatum.

Only a few HD studies employ ML methods with neuroimaging data to differentiate symptom onset from cross-sectional data, as done here. Most of these studies classify HD vs. HC and/or PreHD vs. HC (summarized in supplementary material section S1). However, the separation between HD vs HC or PreHD vs HC has lower clinical translational value since HD diagnosis is effectively done through genetic testing. Therefore, imaging markers are not required to separate HD vs HC or PreHD vs HC.

Moreover, there is a pressing need to better characterise the volumetric changes in the premanifest phase to identify the most suitable participants for therapeutic intervention. What is more difficult but will be more valuable is to examine PreHD vs HD, and even finer-grained (especially PreHD A vs PreHD B) distinction, with a view to predicting those likely to be approaching the clinical motor diagnosis.

In this context, to our knowledge ours is the first study that: (a) employed a novel ensemble ML method to predictively classify HD using neuroimaging data alone; (b) performed fine-grained predictions that span a wide range of the temporal spectrum of HD progression; and (c) used a bigger sample size (n=184) compared to previous studies.^5, 7, 12–14;15–20^

In the following sections we discuss how our results compare to available reported findings, and the strengths, limitations, and future work.

### PreHD vs HD classification

By definition, this distinction is marked by the CMD, i.e., the onset of clinical signs and symptoms. For this reason, the distinction of participants based on sMRI scores has limited practical value and is mostly useful to assess the independent value of MRI at marking CMD. The study by Lavrador^5^ at al., employed SVM classifier and segmented GM from sMRI and FA values from DWI along with several feature selection approaches for classification of HD stage. They utilized specific sub-cortical structures selected a priori—bilateral caudate, putamen and globus pallidus—and investigated each region-of-interest separately by classifying 14 PreHD and 11 early-HD individuals. They reported the highest classification accuracy when using putamen (86.3%±4.2) or caudate (83.0%±3.7), while pallidum (68.1%±5.4) and whole brain (78.2%±6.8) yielded lower accuracy.

Our findings agree with their work indicating that caudate by itself is sufficient for distinguishing between PreHD and HD individuals with good accuracy.^1, 5, 34–37^

### Other finer-grained *binary* classifications

Out of all classification tasks, the stacked model obtained the highest predictive accuracy (max. 76.7%±8.0) and most stable and consistent set of most informative brain regions across repetitions for PreHD A vs PreHD B task. It identified 6 regions of interest: caudate, putamen, occipital, cingulate, lateral ventricles, and sensory motor. These regions largely agree with the reported order of neurodegenerative processes starting from earliest changes in striatal volumes (caudate, putamen) and continuing with cortical regions (occipital, cingulate) and lateral ventricles^21^. Our results also demonstrated that a combination of sMRI features is more accurate and informative in discriminating between progression among premanifest individuals compared to a single striatal volume. These findings emphasize the biomarking potential of multiple brain regions, especially for premanifest stratification and potential applicability to preventive clinical trials.

Like PreHD vs HD classification results, our results showed feature selection instability for PreHD B vs HD1 task. One potential cause of feature instability could be the heterogeneity in these classes, i.e., premanifest classes are defined according to time-to-CMD whilst manifest classes are defined according to the TFC score. Another cause could be if more PreHD B individuals are close to predicted CMD, then their volumetric measures might be quite similar to those in HD1 state.

We also noticed that manifest (HD1 vs HD2) classification accuracy levels were lower compared to other binary classification tasks. However, our results showed that in general a combination of caudate and at least one other brain region (for instance occipital lobe in repeats = 50) could lead to an 8-10% improvement in predictive accuracy compared to accuracy through just caudate. Once more, this finding suggests that disease progression is marked better by combining scores from striatal and non-striatal regions even after symptom manifestation.

### Fine-grained (PreHD A; PreHD B; HD1; HD2) classification

Our stacked model achieved the best predictive accuracy for fine-grained (PreHD A; PreHD B; HD1; HD2) classification. This was the hardest and most complex task wherein the stacked model achieved the maximum accuracy of 55%±6.0, which is low but well above the 25% chance level. The primary reason for the relatively low accuracy could be the heterogeneity in class-definition; each disease state is a priori defined by criteria of different natures that range from CAP score (PreHD A vs. PreHD B) to motor (PreHD B vs. HD1) and functional (HD1 vs. HD2) clinical batteries. Further, we used a dataset containing only 184 individuals, and these are split into four classes that are further divided into CV folds, yielding rather small numbers (9 per fold per class) to train with. Thus, relatively small groups are used to train models that span a vast timescale spanning decades of HD progression.

Although caudate had the highest feature importance since it was never eliminated before the maximal inflection point, using just the caudate led to significantly lower classification accuracy. This finding again points to the distributed biomarking information across multiple brain regions when disease progression is predicted across the full temporal spectrum of the disease using a single cross-sectional MRI scan.

### Brain regions with best combined predictive accuracy for each HD state

Quantifying feature importance enabled us to identify the set of brain regions with the best combined predictive accuracy for distinguishing pairs of HD progression state. These brain regions vary depending on the state but to attain maximum accuracy the stacked model tends to pick at least one brain region from each cluster; for example, one region from the sub-cortical striatal region (caudate), one or more cortical regions (such as occipital lobe or temporal), and one or more from the remaining brain regions (such as lateral ventricles). This indicates that optimal marking of disease progression requires complementary rather than redundant information. Further, it supports the idea that disease progression models should employ multivariate models that capture and exploit the interaction between variables as another source of progression marker. Supporting this view, a recent study in HD by Castro et al.^40^ found similar benefits of multivariate integration and demonstrated that future atrophy in striatal structures is predicted by a deviant correlation between caudate and thalamus.

### Stacked ML model: stable, robust, but does not always improve accuracy

The stacked ensemble model produced a stable and consistent performance for all classification tasks. Its greatest benefit is the model-independent utility, i.e., although the stacked model comprises 6 heterogeneous ML base-models, it seamlessly blended base-model predictions and generated an output that is robust and reliable compared to a monolithic ML model. Compared to the stacked model, we did not find a single base model to systematically peak at all classification tasks, yielding doubts on which of the base models can invariably produce the best result without requiring complex combinations with other models in a stacked approach.

Literature in the ML field has systematically highlighted two main benefits of stacked models; they have a higher prediction accuracy than any contributing model, and they are more stable and robust models.^25, 26^ Here the stacked model matched well the accuracy of the best constituent base model, and mostly improved over it, but it did not result in a systematically improved accuracy for all tasks. One reason for this could be that ensembles are known to improve accuracy when their constituent models are weak and make different errors or disagree on their decisions. However, if base-models are either highly accurate or make similar mistakes, then the meta-model would either have no scope for improvement (in case of former) or will not be able to correct base-model mistakes. Nonetheless, for the most difficult fine-grained task, stacking resulted in significant performance improvement. Altogether, our results lead us to conclude that a stacked model is accurate, more robust than base models, exhibits a graceful performance degradation, and generalizes better than any single ML model.

### Instability in order of feature elimination

Although we found that regions beyond the striatum are required for optimal predictions, the order of brain regions did not stabilize across repeats. Some of the factors contributing to this instability include: **(1) Multicollinearity**: we followed a multivariate approach when investigating the associations between atrophy in different brain regions and HD states. However, the efficiency of such multivariate analysis is subject to correlation among predictive variables. The sMRI measures were highly correlated (see supplementary material section S8 for correlation table) which can lead to the final solution potentially following different paths depending whether variable A or variable B (both highly correlated) is removed first from the model. For example, we noticed that putamen and globus pallidus sometimes swapped places with regard to which one is eliminated first. In the clustering dendrogram, we noticed that these two regions are the most highly correlated, which explains the preference of the selection process to retain only one of the two, but not both. In the bioengineering literature, multicollinearity is known to lead to biased feature importance estimation, loss of predictive accuracy, and reduction of interpretability of the findings.^24^ We considered running dimensionality reduction via principal component analysis prior to modelling disease progression, but ultimately decided to avoid another layer of complexity in the interpretation of results and kept the original brain regions as predictors of progression. Moreover, different brain regions are differentially sensitive to HD-related brain alterations at different disease states and could offer more informative clues than principal factors that dilute their subtle contributions to disease modelling. **(2) Heterogeneous base models**: we noticed that the set of most important features differed depending on ML model.^38^ Therefore, the underlying base-models could “pull” in different directions regarding the importance of each brain region, propagating the instability in the stacked model. **(3) Small sample size**: although we used a relatively large dataset, the amount of signal derived from volumetric anomalies in each subgroup is not sufficient to uncover a clear consistent group of variables among the existing collinearities. Small changes in data or outliers among the various data splits could change the set of extracted relevant features.

The feature selection instabilities highlight an important conclusion in the ongoing HD biomarker development. Most of the literature on MRI markers in HD tends to point to one region (or very few) that is worthy of further development or adoption as an endpoint in clinical trials. Consequently, potential biomarkers are frequently compared for their statistical power and some are found to carry stronger signal. Our study not only identifies multiple regional volumes from across the brain that are informative, but also demonstrates that highly correlated markers can emerge as alternate winners of the feature elimination “race.” A standard study employing only one predictive model without instability analysis could produce a single answer that would be interpreted as the conclusion, and therefore any instability in the model might be concealed. Our findings show that endpoint selection should be thoroughly investigated rather than picked up from a single analysis as is mostly done in traditional research paradigms. Procedures such as bootstrapping, repeated cross-validation or comparative analyses with samples from different studies or geographical regions can help probe the reliability of biomarkers.

### Limitations & future work

We have identified three main limitations of this study. First, we only trained and evaluated our method on the TRACK-HD dataset. We chose this dataset as it has a reasonable balance between premanifest and manifest individuals. However, in the next phase of work we plan to validate our method using datasets such as PREDICT-HD and IMAGE-HD. Second, this research was conducted solely on baseline cross-sectional data. Longitudinal data can provide more accurate estimates of change over time, which can be a better predictor of ongoing disease progression or proximity to the next HD state. While longitudinal analysis is another avenue of application of these models that we plan to test in future studies, the strength of the current model is the classification of individuals based on a single MRI, which can be a realistic scenario for stratifying participants at screening in clinical trials. In this regard, another pressing limitation of this work is the lack of adoption of the more recent HD Integrated Staging System (HD-ISS).^35^ This will be the subject of future extensions of our work.

## Acknowledgements

The authors thank everyone involved in the TRACK-HD study.

## Supplementary material

Supplementary material is available here.

